# COVID-19 PREDICTION IN SOUTH AFRICA: ESTIMATING THE UNASCERTAINED CASES- THE HIDDEN PART OF THE EPIDEMIOLOGICAL ICEBERG

**DOI:** 10.1101/2020.12.10.20247361

**Authors:** Xuelin Gu, Bhramar Mukherjee, Sonali Das, Jyotishka Datta

## Abstract

Understanding the impact of non-pharmaceutical interventions as well as acscounting for the unascertained cases remain critical challenges for epidemiological models for understanding the transmission dynamics of COVID-19 spread. In this paper, we propose a new epidemiological model (eSEIRD) that extends the widely used epidemiological models such as extended Susceptible-Infected-Removed model (eSIR) and SAPHIRE (initially developed and used for analyzing data from Wuhan). We fit these models to the daily ascertained infected (and removed) cases from March 15, 2020 to Dec 31, 2020 in South Africa that reported the largest number of confirmed COVID-19 cases and deaths from the WHO African region. Using the eSEIRD model, the COVID-19 transmission dynamics in South Africa was characterized by the estimated basic reproduction number (*R*_0_) starting at 3.22 (95%CrI: [3.19, 3.23]) then dropping below 2 following a mandatory lockdown implementation and subsequently increasing to 3.27 (95%CrI: [3.27, 3.27]) by the end of 2020. The initial decrease of effective reproduction number followed by an increase suggest the effectiveness of early interventions and the combined effect of relaxing strict interventions and emergence of a new coronavirus variant in South Africa. The low estimated ascertainment rate was found to vary from 1.65% to 9.17% across models and time periods. The overall infection fatality ratio (IFR) was estimated as 0.06% (95%CrI: [0.04%, 0.22%]) accounting for unascertained cases and deaths while the reported case fatality ratio was 2.88% (95% CrI: [2.45%, 6.01%]). The models predict that from December 31, 2020, to April 1, 2021, the predicted cumulative number of infected would reach roughly 70% of total population in South Africa. Besides providing insights on the COVID-19 dynamics in South Africa, we develop powerful forecasting tools that enable estimation of ascertainment rates and IFR while quantifying the effect of intervention measures on COVID-19 spread.

**AMS Classification:** Place Classification here. Leave as is, if there is no classification

## 1 Introduction

The coronavirus disease 2019 (COVID-19) caused by the severe acute respiratory syndrome coronavirus 2 (SARS-CoV-2), was first detected in early December 2019 in Wuhan, China and then quickly spread to majority countries worldwide. At the end of January 2021, over a hundred million people worldwide have been diagnosed with COVID-19 [30], yet the true number of infections in the population remains underestimated, owing to a combination of selection bias from unascertained cases and lack of access to tests early on during the pandemic.

### South Africa

Consider the transmission dynamics we have observed so far in South Africa, the ‘epicenter of the outbreak in the African continent’ [29]. The first case was confirmed in South Africa on March 5, 2020. As of February 23, 2021, there are 1,504,588 confirmed cases of COVID-19 (cumulative total) with 49,150 deaths confirmed in Africa [30]. South Africa remains the worst-hit African country with the largest number of confirmed cases and deaths, by the end of 2020, contributing to 54% of the total confirmed cases and 44% of deaths in the WHO African region, while accounting for only 5% of population [26]. A seroprevalence survey on 4,858 blood donors in South Africa estimated the prevalence of mid January 2021 by province as 63% in the Eastern Cape, 52% in the KwaZulu Natal, 46% in the Free State and 32% in the Northern Cape [38] while the the number of reported cases is 1.83% of the total population at the same time, implying the possibility of a large degree of under-reporting/undetected cases in South Africa. Thus, understanding the key epidemiological constructs for COVID-19 outbreak is paramount for containing the spread of COVID-19 in South Africa, as well as explaining the disparity between seroprevalence estimates and reported number of cases. Two critical factors emerge from analyzing the majority of available evidence of the public health crisis: (1) the unascertained cases and deaths and (2) the role of non-pharmaceutical interventions.

### Unascertained cases and deaths

Based on the clinical characteristics of COVID-19, a majority of patients are symptomatic (roughly 84% according to a recent study [16]), most of whom have mild symptoms [29] and tend to not seek testing and medical care. While private hospitals have reached maximum capacity, public and field hospitals beds have still some margin left with additional challenges due to scarcity of staff [9]. Several recent studies [15, 32, 4] reported that a non-negligible proportion of unascertained cases contributed to the quick spreading of COVID-19. It is suggested that only 1 in 4 mildly ill cases would be detected in South Africa [8]. The relatively lower testing rate in South Africa (Table 1; Figure 1) coupled with a very high test positivity rate especially in July and August [18], suggests inadequacy of testing, as well as the possibility of a large unobserved number of unascertained cases [27]. Thus, modeling both ascertained and unascertained cases and deaths can measure infection fatality ratios (IFRs, the proportion of deaths among all infected individuals [28]) of COVID-19, leading to a better understanding of the clinical severity of the disease.

**Table 1:** Timeline of COVID-19 preventions and interventions in South Africa.

| Date (2020) | Confirmed | Death | Testing rate | Interventions and update |
| --- | --- | --- | --- | --- |
| 5-Mar | 1 | 0 | - | - |
| 10-Mar | 7 | 0 | - | Screening at ports of entry has intensified and escalated. |
| 15-Mar | 51 | 0 | - | Self-quarantine for COVID-19 is recommended. Visas to visitors from high-risk countries (Italy, Iran, South Korea, Spain, Germany, US, UK) are cancelled and previously granted visas are hereby revoked. Gatherings of more than 100 are prohibited. Mass celebrations are canceled. |
| 16-Mar | 62 | 0 | - | Of the 53 land ports, 35 are shut down. |
| 18-Mar | 116 | 0 | - | A travel ban on foreign nationals from high-risk countries such as Germany, US, UK and China. |
| 27-Mar | 1170 | 1 | - | A national lockdown is implemented. Alert level 5 is in effect from midnight 26 March to 30 April. |
| 1-May | 5951 | 116 | 0.004 | A less strict lockdown is in place. Alert level 4 is in effect from 1 to 31 May. Borders will remain closed to international travel, no travel will be allowed between provinces, except for the transport of goods and exceptional circumstances. |
| 1-Jun | 34,357 | 705 | 0.013 | From 1 June 2020 alert level 3 will be in effect. Restrictions on many activities, including at workplaces and socially, to address a high risk of transmission. |
| 18-Aug | 592,106 | 12,264 | 0.059 | Alert level 2 is in effect. |
| 21-Sep | 661,898 | 15,992 | 0.07 | Alert level 1 is in effect. |

**Figure 1:**
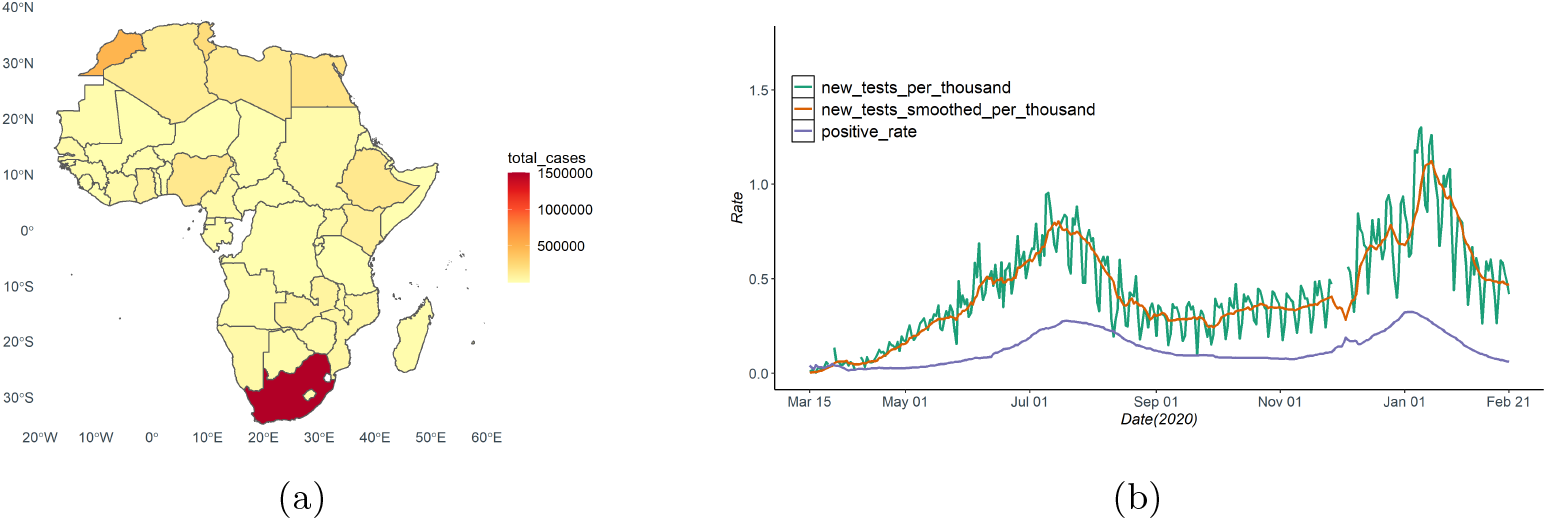
(a) Total cases by country in the African continent; (b) The 7-day average testing positive rate of COVID-19 in South Africa during the study period.

### Interventions

With a universal goal to ‘flatten the curve’, a series of non-pharmaceutical interventions were implemented by the government in South Africa, that have been gradually lifted since early May 2020 [25]. On March 27, 2020, South Africa adopted a three-week nationwide hard-lockdown (level 5) along with closure of its international borders, which was extended to April 30, 2020. Thereafter, to balance the positive health effects of strict interventions against their economic costs [1], South Africa began a gradual and phased recovery of economic activities with the lockdown restriction eased to level 4 [25], allowing inter-provincial travel only for essential services. From June 1, national restrictions were lowered to level 3 allowing for inter-provincial travel and school opening and eased to level 2 and level 1 from August 18 and September 21, 2020 (Table 1). Face-mask wearing was mandatory in public places at all times, with limitations on gatherings, and sale of alcohol and cigarettes were restricted [12]. Although these interventions implemented at an early stage had a higher potential for pandemic containment, previous studies [12, 24, 42] reported a consistently large value for the estimated basic reproduction number (*R*_0_) ranging from 2.2 to 3.2 in South Africa by models trained with data in relatively early time windows. Using data observed under various intervention scenarios over a longer period of time, we carry out a thorough investigation to assess the current COVID-19 spread and the effect of these interventions, which will provide valuable insights into the transition dynamics of COVID-19 and intervention deployment in South Africa, and beyond.

### Epidemiological models

Since the early days of the pandemic, researchers have responded to the unprecedented public health crisis by providing forecasts and alternative scenarios to inform decision-making, both locally and globally. This has resulted in hundreds of mathematical models of varying complexity. The Susceptible-Infectious-Removed or SIR model [20] is arguably the most commonly used epidemiological models for modeling the trajectory of an infectious disease. A recent extension of SIR, called extended-SIR or eSIR [35], was developed to incorporate user-specified non-pharmaceutical interventions and quarantine protocols into a Bayesian hierarchical Beta-Dirichlet state-space model, which was successfully applied to model COVID-19 dynamics in India [33]. One major advantage of this Bayesian hierarchical structure is that uncertainty associated with all parameters and functions of parameters can be calculated from posterior draws without relying on large-sample approximations [33]. Extending the simple compartment structure in eSIR model, the SAPHIRE model [40] delineated the full transmission COVID-19 dynamics in Wuhan, China with additional compartments by introducing unobserved categories [15].

In this article, we extended the eSIR approach to the eSEIRD model to combine the advantages of the two existing models, using a Bayesian hierarchical structure to introduce additional unobserved compartments and characterize uncertainty in critical epidemiological parameters including basic reproduction number, ascertainment rate and IFR, with input data as observed counts for cases, recoveries and deaths. Furthermore, we applied these three models and compared the results of the eSEIRD model with two of the existing alter-natives, namely the eSIR and the SAPHIRE model, with the following primary objectives: (i) characterizing the COVID-19 dynamics from March 15 to December 31, 2020 – under different time-varying intervention scenarios; (ii) evaluating the effectiveness of the main non-pharmaceutical interventions such as lockdown, and mandatory wearing of face-mask in public places; (iii) capturing the uncertainty in estimating the ascertainment rate and IFR; and (iv) forecasting the future of COVID-19 spread in South Africa.

The organization of this paper is as follows: we describe the two competing epidemiological models and our proposed extension in §2. The study design and parameter settings for modeling COVID-19 transmission in South Africa are described in §3, and the results and their possible implications are described in §4. We conclude with a discussion of nuances and limitation of the methods and sources of data used here and suggest future directions in §5.

### 2 Statistical Methodology

We propose an extension of the eSIR model, called eSEIRD, and compare it against two existing epidemiological methods, the eSIR and SAPHIRE model. In this section, we describe the dynamic systems and the hierarchical models underlying these three epidemiological models. The schematic diagrams for the three compartmental models are shown in Fig. 2 (eSIR and SAPHIRE) and Fig 3 (eSEIRD).

**Figure 2:**
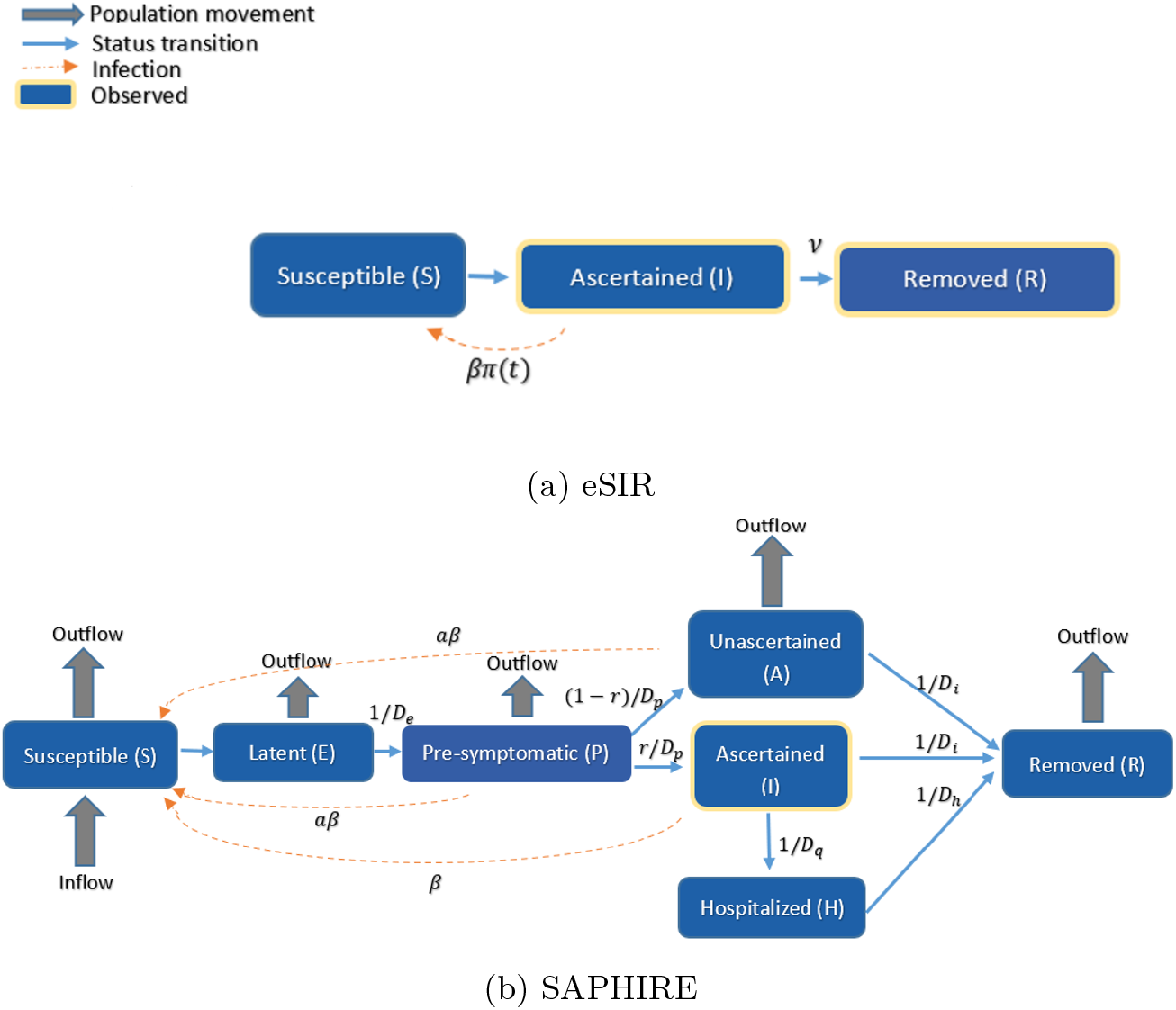
Schematic diagram of the two models (a) eSIR; (b) SAPHIRE.

**Figure 3:**
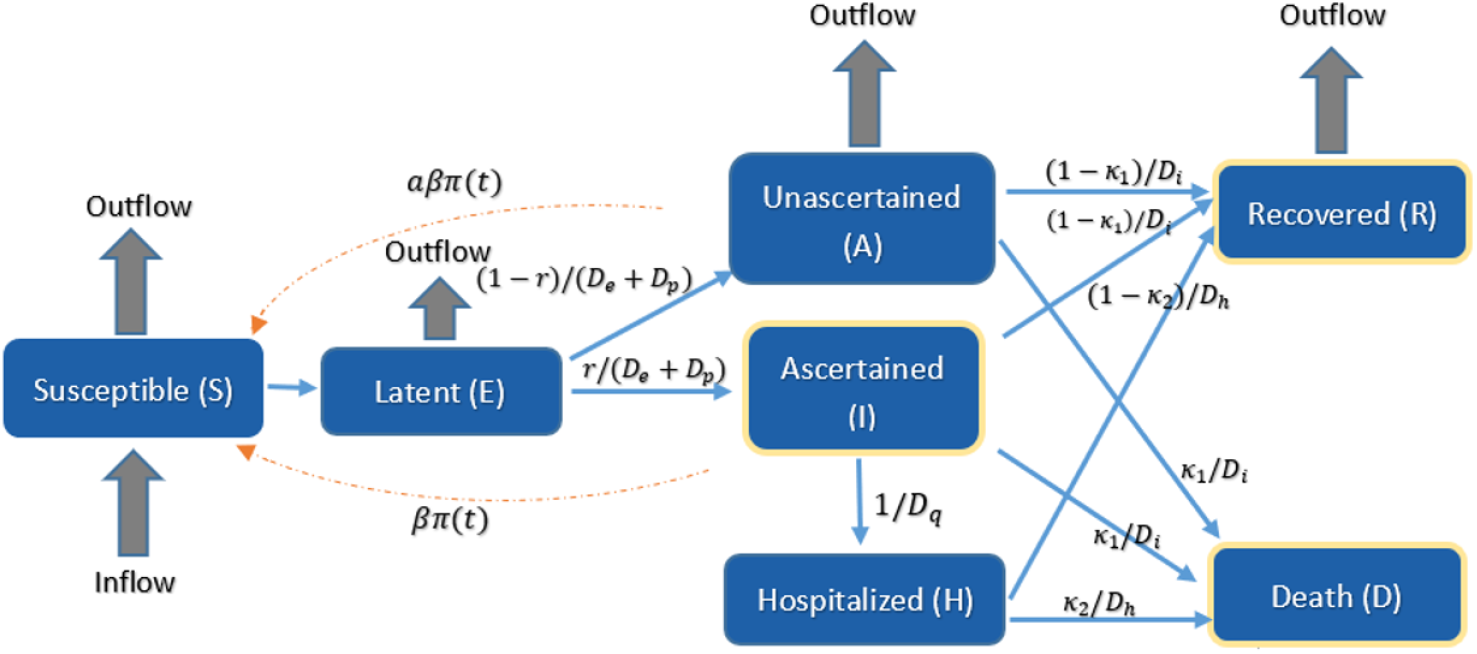
Schematic diagram of the proposed eSEIRD model

### 2.1 eSIR model

The eSIR model assumes the true underlying probabilities of the three compartments susceptible (S), infectious (I) and removed (R) follow a latent Markov transition process and require observed daily proportions of cumulative infected and removed cases as input [31, 35]. The observed proportions of infected and removed cases on day *t* are denoted by 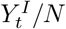 and 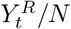 (the infected and removed counts 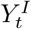 and 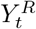 divided by total population size *N*) respectively. Further, we denote the true underlying probabilities of the three compartments on day t by 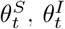 and 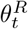, respectively, and assume that for any *t*, 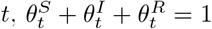 The following set of differential equations describe the dynamic system for the usual SIR model on the true proportions.

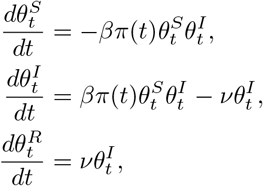

where *β >* 0 denotes the disease transmission rate, and *ν >* 0 denotes the removal rate (see Fig. 2(a) for a schematic representation). The basic reproduction number *R*_0_ = *β/γ* indicates the expected number of cases generated by one infected case in the absence of any intervention and assuming that the whole population is susceptible.

### Hierarchical model

The eSIR model works by assuming that two observed time series of daily proportions of infected and removed cases are emitted from two Beta-Dirichlet state-space model, independent conditionally on the underlying process governed by the Markov SIR process:

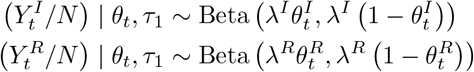

and the Markov process associated with the latent proportions is built as:

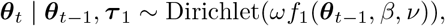

where, ***θ***_*t*_ denotes the vector the true underlying probabilities of the compartments on day *t* whose mean is modeled as an unknown function of the probability vector from the previous time point, along with the transition parameters; 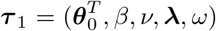. The function *f*_1_(·) is then solved as the mean transition probability determined by the SIR dynamic system, using a fourth order Runge-Kutta (RK4) approximation (see supplementary §S.3 for the solution):

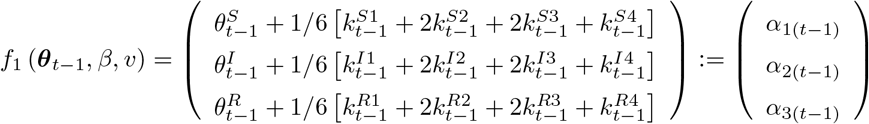

Computational details for the eSIR model such as posterior sampling strategy using MCMC algorithm is complemented by the R package publicly available at https://github.com/lilywang1988/eSIR.

### 2.2 SAPHIRE model

The SAPHIRE model [15] is an extension of the basic SIR model with additional compartments to allow for unobserved categories such as ascertained, unascertained and hospitalized population. Specifically, in a SAPHIRE model the population is compartmentalized into susceptible (S), exposed (E), presymptomatic infectious (P), ascertained infectious (I), unascertained infectious (A), isolation in hospital (H) and removed (R). Denoting the true underlying accounts of the S, E, P, A, I, H and R compartments on day *t* by *S*_*t*_, *E*_*t*_, *P*_*t*_, *A*_*t*_, *I*_*t*_, *H*_*t*_ and *R*_*t*_, respectively, the dynamics of these compartments across time *t* were described by the following set of differential equations (see Fig. 2(b) for a schematic representation):

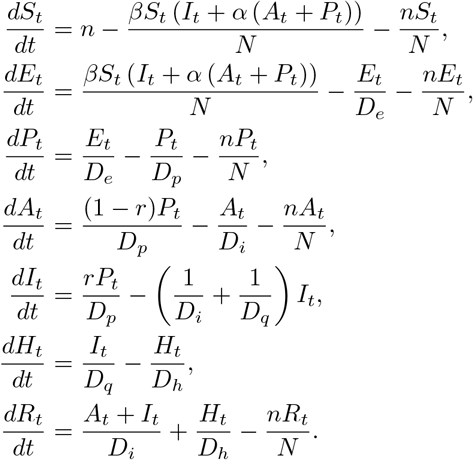

To fit the SAPHIRE model, the observed number of ascertained cases in which individu-als experienced symptom onset on day *t*, 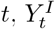, were assumed to follow a Poisson distribution:

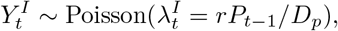

where *P*_*t−* 1_ denotes the underlying number of pre-symptomatic individuals and r denotes the ascertained rate. For an observation window spanning *t* = 1 to *t* = *T*, the sampling pseudo-likelihood function for the underlying prevalence parameters is given by:

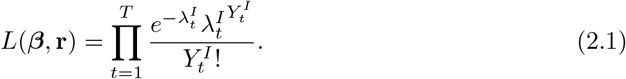

after plugging 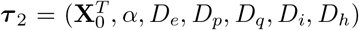 and **X**_*t*_ = (*S*_*t*_, *E*_*t*_, *P*_*t*_, *A*_*t*_, *I*_*t*_, *H*_*t*_, *R*_*t*_). Here, *X*_*t*_ denotes the vector of the underlying population counts of the compartments at time *t*. With the initial values for the compartments **X**_0_ set at pre-fixed values, the pseudo-likelihood function in (2.1) can be approximated as a function of the parameters of interest, i.e. ***β*** and *r*, by the following steps:

**Step 0** The transition parameters 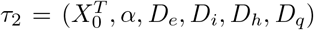 and the initial values for the compartments *X*_0_ = (*S*_0_, *E*_0_, *P*_0_, *A*_0_, *I*_0_, *H*_0_, *R*_0_) are fixed;

**Step 1** Use the differential equations to generate the change of each compartment at time *t* = 1, i.e. *dX*_*t*_*/dt* = (*dS*_*t*_*/dt, dE*_*t*_*/dt, dP*_*t*_*/dt, dA*_*t*_*/dt, dI*_*t*_*/dt, dH*_*t*_*/dt, dR*_*t*_*/dt*);

**Step 2** Compute the state values of each compartment at time 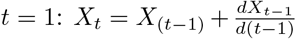, and the expected new ascertained cases 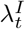 on day *t*;

**Step 3** Repeat the step 1-3 for *t* = 2, 3, 4, …, *T*.

For the SAPHIRE model, the MCMC algorithm is implemented with the delayed rejection adaptive metropolis algorithm implemented in the R package BayesianTools (version 0.1.7) to collect posterior samples of the underlying parameters *r* and *β*, then calculate the derived quantities the effective reproduction number *R*_*e*_ based on the posterior draws. We refer the reader to [15] for more details.

### 2.3 eSEIRD model

Similar to the hierarchical structure used in eSIR model, this eSEIRD model (*vide* Fig. 3) works by assuming that the true underlying probabilities of the 7 compartments follow a latent Markov transition process which fits not only the count of daily infected, but also the recovered and death counts.

The dynamics of these 7 compartments across time *t* were described by the following set of ordinary differential equations:

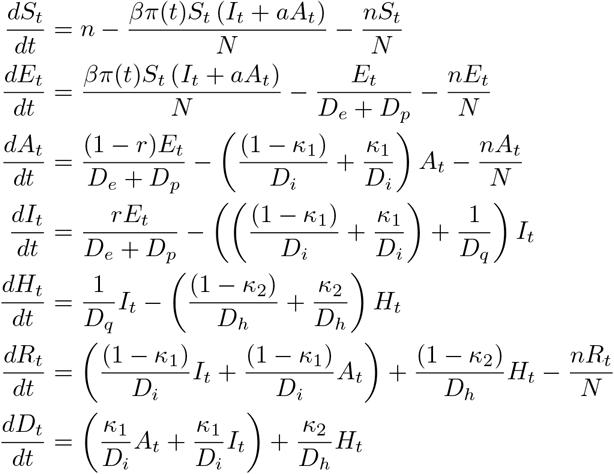

### Hierarchical model

We assumed three observed time series of daily counts of infected, recovered and death cases are emitted from Poisson state-space models, independent conditionally on the underlying process, and the Markov process associated with the latent proportions is constructed as:

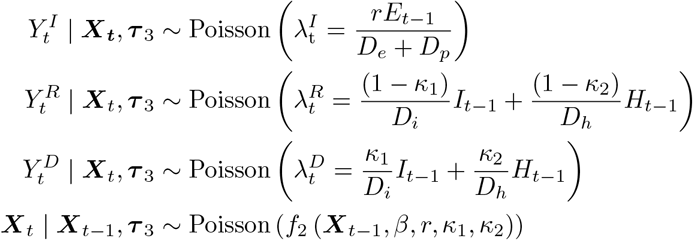

where *X*_*t*_ = (*S*_*t*_, *E*_*t*_, *A*_*t*_, *I*_*t*_, *H*_*t*_, *R*_*t*_, *D*_*t*_) denotes the vector of the underlying population counts of the 7 compartments; 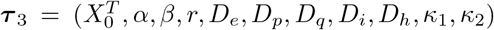 denotes the whole set of parameters where 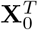denotes the prior for the initial states and *κ*_1_ and *κ*_2_ denote the IFR for non-hospitalized and hospitalized cases, respectively. The function *f*_2_(·) is also solved using RK4 approximation as before (see the solution in Supplementary §S.4).

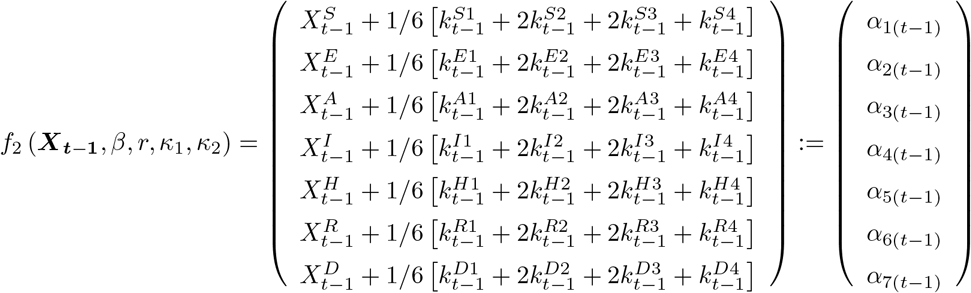

We implemented the MCMC algorithm to sample from the posterior distribution of the underlying parameters *r* and *β*, and calculate the derived quantities:

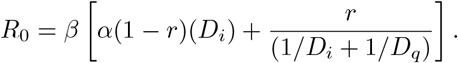

We obtain the posterior mean estimates and credible intervals for the unknown parameters in the model. Because of the hierarchical structure in the state-space model considered in this model, the posterior sampling can be done in a straightforward fashion like eSIR using the R package rjags.

## 3 Modeling COVID-19 transmission dynamics in South Africa

### 3.1 Study Design and Data Source

COVID-19 daily time series data for South Africa were extracted from the COVID-19 Data Repository by the Center for Systems Science and Engineering (CSSE) at Johns Hopkins University [39] from the onset of the first 50 confirmed case (March 15, 2020) to February 21, 2021. We fitted the models using data up to December 31, 2020 and predicted the state of COVID-19 infection in South Africa in a time window, from January 1 to April 1, 2021. To compare the model prediction performance of different models, we used the symmetric mean absolute percentage error (SMAPE), given by:

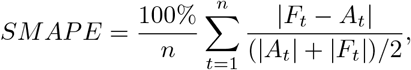

where *A*_*t*_ is the observed value and *F*_*t*_ is the forecast value in the same time period. This design enabled us to select an optimal modeling strategy for South Africa data and check the robustness of prediction performance across different models.

#### 3.1.1 Prior Specification

We describe the prior choices and where appropriate, initial values for the model hyper-parameters in this section and a complete summary and list of notations and assumptions are available in Supplementary Table S.1.1. To begin with, we assumed a constant population size (*N* = 57, 779, 622) for all models and fixed a few transition parameters below in the SAPHIRE and eSEIRD model. First, we set an equal number of daily inbound and outbound travelers (*n*), in which *n* = 4 *×* 10^*−*4^ *N* from March 15 to 25, 2020 estimated by the number of international travelers to South Africa in 2018[3], otherwise *n* = 0 when border closed, i.e. after March 26. We fixed the transmissibility ratio between unascertained and ascertained cases at *α* = 0.55 assuming lower transmissibility for unascertained cases [2], an incubation period of 5.2 days, and a pre-symptomatic infectious period of *D*_*p*_ = 2.3 days [23, 17], implying a latent period of *D*_*e*_ = 2.9 days. The mean of total infectious period was *D*_*i*_ + *D*_*p*_ = 5.2 days [23], assuming constant infectiousness across the pre-symptomatic and symptomatic phases of ascertained cases [22], thus, the mean symptomatic infectious period was *D*_*i*_ = 2.9 days. We set the period of ascertained cases from reporting to hospitalization *D*_*q*_ = 7 days, the same as the median interval from symptom onset to admission reported [10, 13]. The period from being admitted in hospital to discharge or death was assumed as *D*_*h*_ = 8.6 days [41]. We fit the SAPHIRE and eSEIRD model in six time periods of 2020: March 15-March 26, March 27-April 30, May 1-May 31, June 1-August 17, August 18-September 20, and September 21-December 31, separated by the change-points of the lockdown strictness level, and denote the ascertained rate and transmission rate in the time periods as *r*_1_, *r*_2_, *r*_3_, *r*_4_, *r*_5_,*r*_6_, *β*_1_, *β*_2_, *β*_3_, *β*_4_, *β*_5_ and *β*_6_. In addition, we denote the IFR for non-hospitalized cases *κ*_11_, *κ*_12_, *κ*_13_, *κ*_14_, *κ*_15_, *κ*_16_ and for hospitalized cases *κ*_21_, *κ*_22_, *κ*_23_, *κ*_24_, *κ*_25_, *κ*_26_ in eSEIRD model.

### Choice of Initial states

For the eSIR model, the prior mean for the initial infected/removed proportion was set at the observed infected/removed proportion on March 15, 2020, and that for the susceptible proportion was the total number of the population minus the infected and removed proportions [35]. For the SAPHIRE model, other than setting prior parameters for initial states, we set the number of initial latent cases *E*(0) was the sum of those ascertained and unascertained cases with onset during March 15-17, 2020 as *D*_*e*_ = 2.9 days [15] and the number of initial pre-symptomatic cases *P* (0) was that from March 18-19, 2020 as *D*_*p*_ = 2.3 days [15]. The number of ascertained symptomatic cases *I*(0) was assumed as the number of observed infected cases on March 15, 2020 excluding *H*(0), *R*(0) and *D*(0) (the initial numbers for hospitalized, recovered, and deaths). The initial ascertainment rate (*r*_0_) was assumed as 0.10 as reported in literature [4, 6], implying *A*(0) = 0.90*/*0.10*I*(0), and a sensitivity analysis with *r*_0_ = 0.25 was conducted to address weak information for *r*_0_ obtained in South Africa and variation of *r*_0_ in different scenarios. *H*(0) was assumed as 50% of the observed ascertained cases on March 9, 2020 (by assuming the period from reported to hospitalized was 7 days [10, 13] at the early stage of the pandemic). In addition, we denoted *R*(0) as the sum of observed recovered and death cases on March 15. The number of initial susceptible cases *S*(0) was calculated as the total population (*N*) minus *E*(0), *P* (0), *I*(0), *A*(0) and *R*(0).

In the eSEIRD model, we set the prior mean of initial ascertained, unascertained and hospitalized cases as *I*(0), *A*(0) and *H*(0) discussed above. However, since the latent compartment incorporates the pre-symptomatic cases, the mean of the initial latent cases was set as the sum of those ascertained and unascertained cases with onset during March 15-19, 2020 as *D*_*e*_ + *D*_*p*_ = 5.2 days [15].The prior mean of initial recoveries and deaths were fixed as the number of observed recovered and death cases on March 15, 2020, respectively. Therefore, the prior mean of initial susceptible compartment was set as the total population excluding the mean of other compartments.

### Prior distributions

In the eSIR model, the log-normal priors were used for the removed rate *ν* and the basic reproduction number *R*_0_, in particular *ν ∼ LogN* (2.955, 0.910),with *E*(*ν*) = 0.082 and *SD*(*ν*) = 0.1 [35], and *R*_0_(= *β/ν*) *∼ LogN* (0.582, 0.223) with *E*(*R*_0_) = 3.2 and *SD*(*R*_0_) = 1 [33]. Flat Gamma priors were used for the scale parameters of the Beta-Dirichlet distributions as follows [33]:

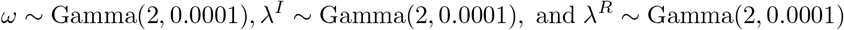

In the eSEIRD model, for the six time periods, all the transmission rates *β*_1_, *β*_2_, *β*_3_, *β*_4_, *β*_5_ and *β*_6_ were given a *U* (0, 2) prior, ascertained rates *r*_1_, *r*_2_, *r*_3_, *r*_4_, *r*_5_ and *r*_6_ were given Beta(10, 90) prior [21], the IFR for non-hospitalized cases *κ*_11_, *κ*_12_, *κ*_13_, *κ*_14_, *κ*_15_, *κ*_16_ *∼* Beta(0.03, 2.93) and for hospitalized cases *κ*_21_, *κ*_22_, *κ*_23_, *κ*_24_, *κ*_25_, *κ*_26_ *∼* Beta(0.44, 1.76) with mean equal to 0.1% and 20%, respectively [41].

In addition, to account for the effect of time-varying contact rate during the prediction period, we set a time-varying contact rate modifier *π*(*t*) in the eSIR and eSEIRD model where *t* from January 1 to April 1, 2021: *π*(*t*) was set as 0.75 since the lockdown was tuned to level 3 after December 28, 2020. Note that the modifier *π*(*t*) is a conjectural quantity and hence must be guided by empirical studies [33]. Using MCMC sampling method for the eSIR and eSEIRD model, we set the adaptation number to be 10^4^, thinned by 10 draws to reduce auto-correlation, and set a burn-in period of 5 *×* 10^4^ draws under 10^5^ iterations for 4 parallel chains.

We fit the SAPHIRE model in six time periods as in the eSEIRD model. We used *r*_1_ *∼ Beta*(10, 90) and reparameterized *r*_2_, *r*_3_, *r*_4_, *r*_5_, and *r*_6_ by

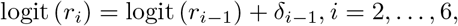

where logit(*r*) = log(*r/*(1*−r*)), and *δ*_*i*_ *∼ 𝒩* (0, 1), for *i* = 1, …, 5. We use non-informative prior distributions for transmission rates *β*_*i*_ *∼ 𝒰* (0, 2) for *i* = 1, …, 6 to reflect lack of information about these hyperparameters [15]. Therefore, *β* and *r* were assumed to follow different distributions for these six time periods. Finally, the effective reproduction number can be derived to be:

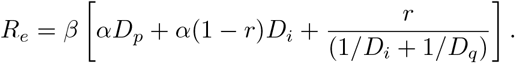

We set a burn-in period of 10^5^ iterations and continued to run 10^5^ iterations with a sampling step size of 10 iterations for the MCMC algorithm of the SAPHIRE model.

A comparison between assumptions of the three models in the Supplementary §S.2. All analyses were conducted in R (version 4.0.0), and source codes are available at https://github.com/umich-cphds/south_africa_modeling. Posterior mean and corresponding 95% credible interval (95% CrI) were reported for the parameters of interests.

## 4 Results

Here we present the detailed results for South Africa, subdivided into estimation of key epidemiological parameters, short-term and long-term forecasts, and finally model evaluation in terms prediction and quality of fit.

### 4.1 Reproduction number and intervention evaluation

The estimated posterior mean of *R*_0_ was 1.18 (95%CrI: [1.09, 1.28]) in the eSIR model throughout the training period while in the eSEIRD model, the value of *R*_0_ started at 3.22 (95%CrI: [3.19, 3.23]) then dropped though still significantly above 1 after the lockdown implementation and increased to 3.27 (95%CrI: [3.27, 3.27]) during the last three months of 2020 (Table 2). It suggests that the effective contact rate decreased by more than 50% over the lockdown time period and attained the lowest point in its trajectory during August to September, 2020 though the lockdown was eased to a relatively less strict level. On the other hand, the effective reproduction number (*R*_*e*_) in different lockdown periods estimated by the SAPHIRE model demonstrates that a similar trend but the magnitude of the estimated *R*_*e*_ decreased dramatically when *r*_0_ increases from 0.10 to 0.25 (Table 2; Fig. 4), possibly suggesting lack of robustness with respect to the choice of initial *r*_0_.

**Table 2:**
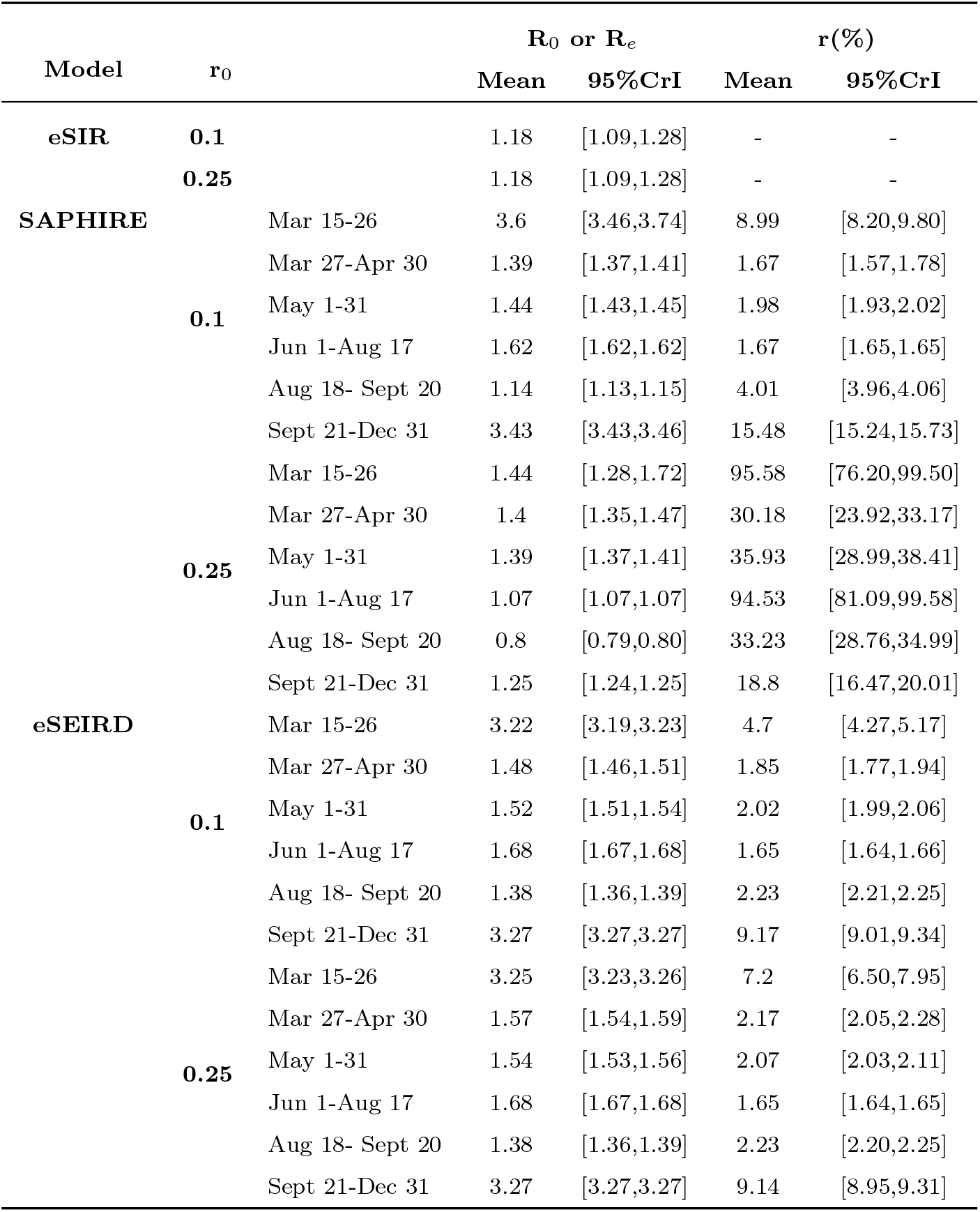
The posterior mean and credible intervals of the basic/effective reproduction number (*R*_0_ or *R*_*e*_) and ascertained rate (*r*) obtained from different models and settings.

**Figure 4:**
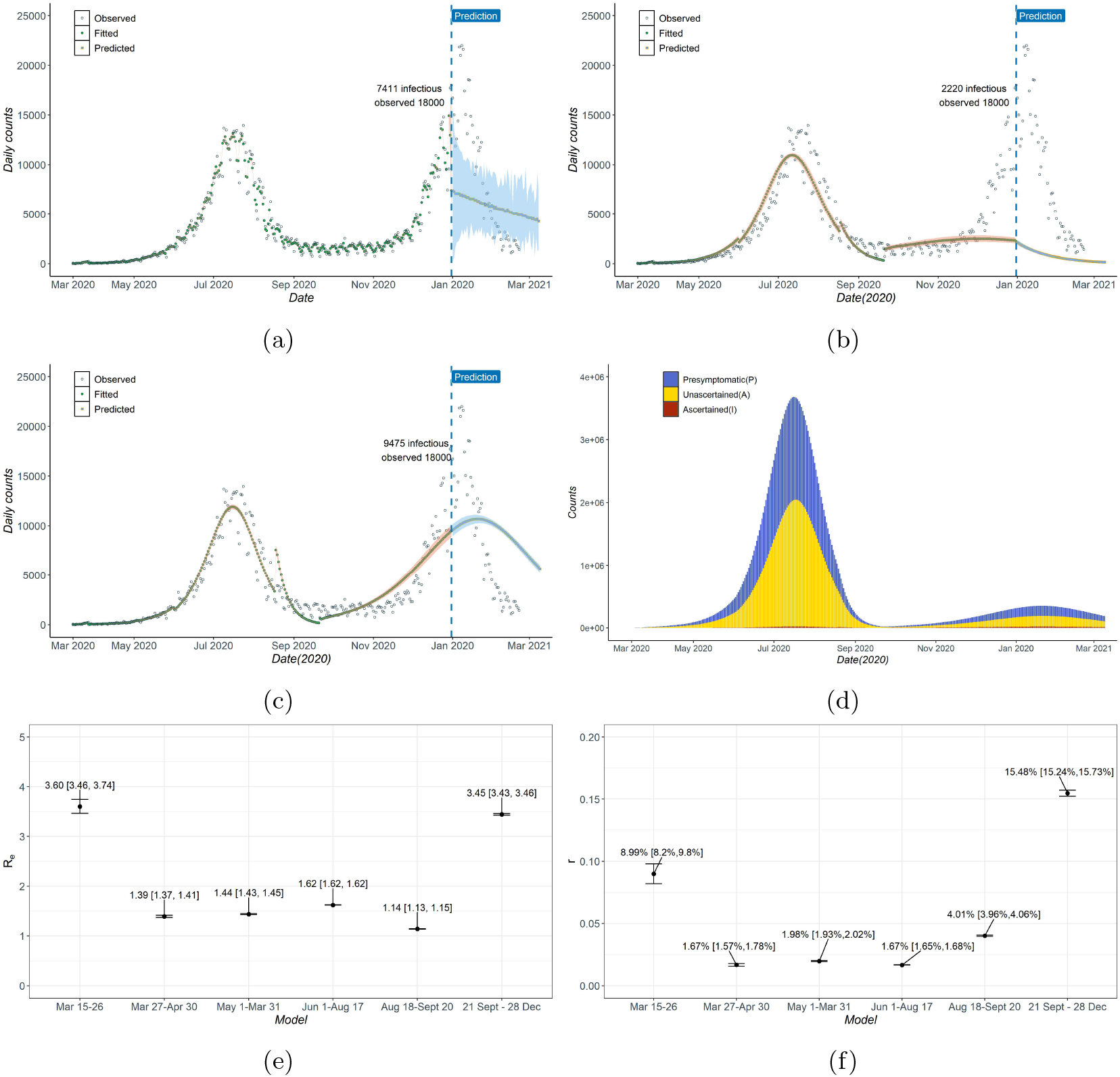
(a)-(c) Daily new number of ascertained infections cases estimated by the models compared with observed data: (a) eSIR, (b) SAPHIRE, and (c) eSEIRD; (d) Current pre-symptomatic/unascertained/ascertained infectious in the SAPHIRE model; (e)-(f) Estimated effective reproduction number (*R*_*e*_) and ascertained rate (r) in the SAPHIRE model in four time periods. (Assume initial ascertained rate (*r*_0_) equal to 0.10.)

### 4.2 Short-term and long-term forecasts

We forecast the total cumulative number of infections, including unascertained cases, in the SAPHIRE model up to February 28, 2021 depending on the time-period considered for estimating the trend. The estimated cumulative number of infections are: (a) 43.3 million if the trend of the least strict lockdown (level 1) was assumed, (b) 36.9-37.8 million if the trend of the lockdown level 2-4 was assumed, and (c) 29.3 million if the trend of strict lockdown was assumed (*r*_0_ = 0.10). However, the short-term forecasts in SAPHIRE model exhibits lack of robustness under different *r*_0_ settings, for example, when *r*_0_ = 0.25 the estimated cumulative number of infections was 10.6 million if the trend of the least strict lockdown (level 1) was assumed and 0.9 million if the trend of the lockdown level 2 was assumed. In the eSEIRD model, the predicted total cumulative number of cases reach 41.2 million under *r*_0_ = 0.10, and 41.6 million under *r*_0_ = 0.25, along with estimated total deaths counts as 35 or 37 thousand when *r*_0_ = 0.10 or 0.25, respectively, by February 28, 2021. Furthermore, we used the eSEIRD model to forecast the epidemic trajectory for a relatively longer time period, where we found that by April 1, the cumulative number of total infected and deaths would reach roughly 41.3 million (which is around 70% of the total population in South Africa) and 35 thousand, respectively.

### 4.3 Fitting and prediction performance

All the three candidate models were applied to the COVID-19 data in South Africa with high accuracy as the estimated daily new cases were close to the observed numbers from March to October, 2020 (Fig. 4 (a)-(c)). However, the eSEIRD model showed a poorer fit during the second pandemic wave in South Africa from November to December 2020, compared to the other two models. The eSIR model performed best in terms of fitting the cumulative ascertained cases with the smallest SMAPE (2.43% when *r*_0_=0.10) while the SAPHIRE model had the second smallest training SMAPE (Table 4). In terms of predictive accuracy, the SAPHIRE model performed best with the smallest SMAPE (4.41% for 15 days and 5.92% for 31 days when *r*_0_=0.10) while the eSIR model had the second smallest SMAPE (6.90% for 15 days and 10.78% for 31 days when *r*_0_=0.10) (Table 4). We note that for a few selected important time points, the estimated number of cumulative ascertained infected cases for the eSEIRD model was closest to the observed on December 31, 2020, while the predicted number of cases in the eSIR and SAPHIRE model are closer to the observed on January 31, 2021 (Table 3). The predictive performances for the three competing models substantiate their credibility in terms of capturing the transmission dynamics for the time-period considered in this study.

**Table 3:** Comparison of the models regarding the cumulative ascertained infected and death with the observed (in thousands). Bold-faced entries indicate column winners regarding the closeness to the observed.

| Model | $r_0$ | Infected | | | Death | | |
| --- | --- | --- | --- | --- | --- | --- | --- |
|  |  | Estimation | Prediction |  | Estimation | Prediction |  |
|  |  | 31-Dec | Jan 31 | 28-Feb | 31-Dec | 31-Jan | 28-Feb |
|  |  | 2020 | 2021 | 2021 | 2020 | 2021 | 2021 |
| eSIR | 0.1 | 1052 | 1256 | 1403 | - | - | - |
|  | 0.25 | 1052 | **1256** | 1402 | - | - | - |
| SAPHIRE | 0.1 | 1058 | **1379** | 1624 | - | - | - |
|  | 0.25 | 1055 | 1576 | 2320 | - | - | - |
| eSEIRD | 0.1 | **878** | 917 | 928 | 31 | 34 | 35 |
|  | 0.25 | **924** | 966 | 977 | 33 | 36 | 37 |
| Observed | - | 931 | 1381 | - | 25 | 40 | - |

**Table 4:**
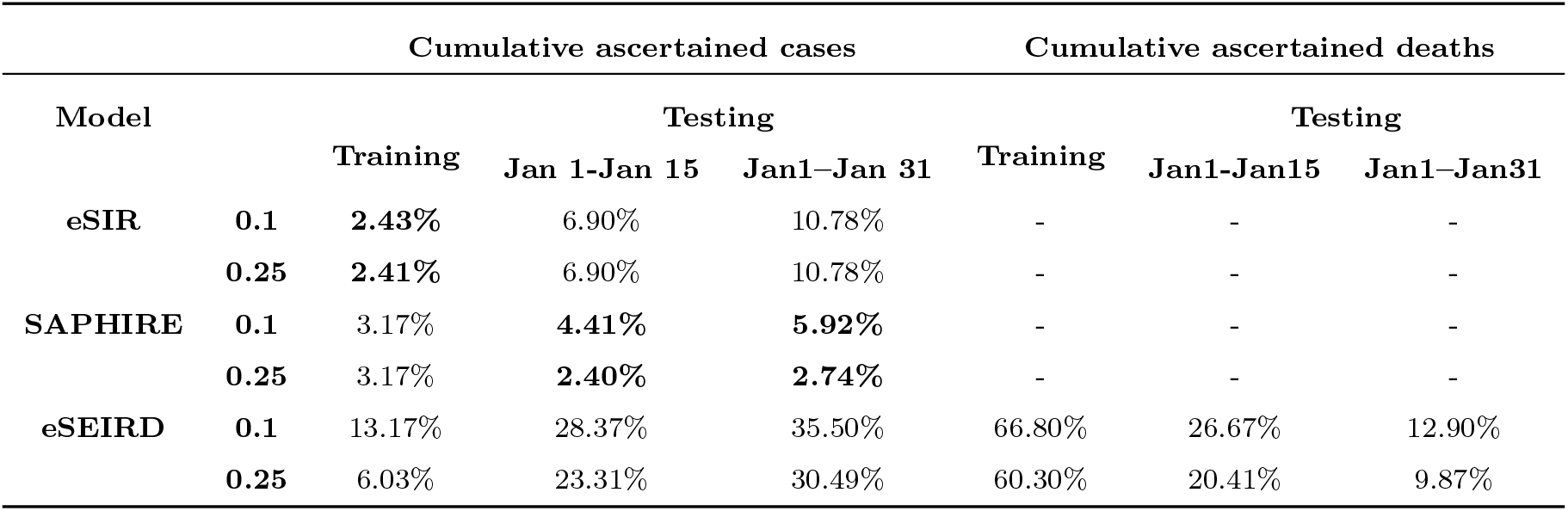
Symmetric mean absolute percentage error (SMAPE) of short-term forecasting. Bold-faced entries indicate column winners regarding prediction performance.

### 4.4 Unascertained cases and deaths

As demonstrated by SAPHIRE modeling results in Figure 4 (d), the large number of unascertained and pre-symptomatic cases contributed to the rapid spread of disease.The estimated ascertained rates were very low, starting at 8.99% (95% CrI: [8.20%, 9.80%]), decreasing to below 2 during level 5 to level 3 lockdown and then increasing to 15.48% (95% CrI: [15.24%, 15.73%]) during the second pandemic wave in South Africa, respectively (Table 2; Fig. 4 (f)). Similarly, in the eSEIRD model, the estimated ascertained rates were also at a very low level (1.65% to 9.17%) and had a similar trend as in SAPHIRE model (*vide* Table 2). As mentioned before, the estimated ascertained rates were robust with respect to choices for *r*_0_ in eSEIRD model, but changes drastically in SAPHIRE model with *r*_0_ changed to 0.25: 95.58% (95%CrI: [76.20%, 99.50%]) before lockdown, between 30.18% to 35.93% during level 5 and 4 lockdown, 95.53% (95%CrI: [81.09%, 99.58%]) in level 3 lockdown, and then decreasing form 33.23% to 18.80% in level 2 to level 1 lockdown.

By the eSEIRD model, the overall IFR was estimated as 0.06% (95%CrI: [0.04%, 0.22%]) throughout the whole time period taking the reported and unreported cases and deaths into account while the observed overall case fatality ratio was estimated as 2.88% (95% CrI: [2.45%, 6.01%]) (Fig. 5). Furthermore, the eSEIRD model provided Bayesian estimates for IFR and deaths among hospitalized and non-hospitalized cases. The estimated IFR for hospitalized cases was 15.28% (95%CrI: [0.01%, 69.10%]) before lockdown and increased to 65.86% (95%CrI: [51.00%, 82.91%]) in the first time period of lockdown. After that, the IFR for hospitalized cases decreased from 22.9% (95%CrI: [20.75%, 25.18%]) to 7.46% (95%CrI: [7.46%, 7.71%]) during May to September. By the end of 2020, it again increased to 19.25% (95%CrI: [18.82%, 19.69%]). The IFR of hospitalized cases was much larger than that of non-hospitalized cases (less than 0.01%),and these estimates were robust to the choice of *r*_0_.

**Figure 5:**
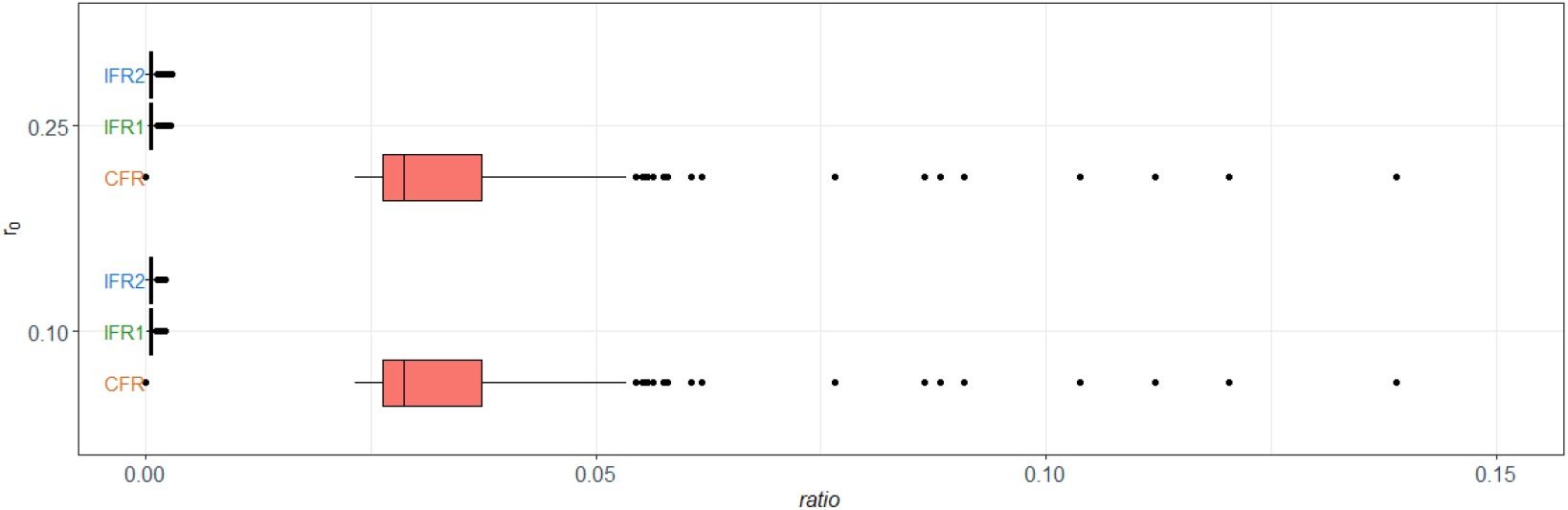
Case fatality ratio (CFR) and estimated infection fatality ratio (IFR) in the eSEIRD model. CFR =(Number of reported deaths)/(Number of reported deaths and recovered) ; IFR1 = (Number of reported deaths)/(Number of of reported and unreported cases); and IFR2 = (Number of reported and unreported deaths)/(Number of of reported and unreported cases)[20].

## 5 Discussion

In this paper, we propose a new infectious disease forecasting model that incorporates the unascertained cases, population movement over different time periods, and the effect of intervention strategies in a unified way and use it to investigate the spread of COVID-19 in South Africa, the hardest hit country on the African continent. The methodological tools developed here can be used to estimate the IFR as well as estimate actual COVID-19 deaths from the reported death counts.

The lockdown intervention and mandatory face-mask wearing in public places employed in South Africa seemed to contain the spread of COVID-19 effectively as the *R*_*e*_ decreased dramatically initially but increased later following the relaxation of lockdown stringency afterwards. However, the *R*_*e*_ was consistently above 1 throughout the whole period analyzed, which implies the interventions failed to dampen the transmission fully, further substantiated by the basic reproduction number estimates in the eSEIRD model as well. This agrees with the public health experts advice of carefully implemented intervention policies while taking account their potential economic costs [37].

We also found that the estimated ascertainment rate is very low in South Africa compared to that reported for many other countries [15, 4, 32], also implied by the low testing rate and high testing positive rate in South Africa. As of February 21, the number of total tests conducted is 8.9 million, suggesting that about 15.4% population were tested [18].

Furthermore, the estimated ascertainment rate is consistent with that in other multiple global epicenters under severe pandemic of COVID-19, such as France, the United States, Italy and Spain in March [21]. The large number of unascertained cases is likely to contribute significantly to the continuing spread of COVID-19 [23, 7, 19]. Our findings suggest that there are around 70% of the total population in South Africa infected by December 31, 2020, which is roughly consistent with the seroprevalence survey conducted in South Africa that the estimated prevalence is around 63% in the Eastern Cape, one of the pandemic centers in South Africa [38]. Despite the potential high prevalence of COVID, the second wave of pandemic appeared in South Africa and other pandemic centers like Brazil, which may due to the waning immunity against infection with the time to the first wave increasing, and the coronavirus lineages might have higher inherent transmissibility than the pre-existing lineages and be able to evade the immunity generated in response to previous infection [34]. To prevent potential resurgence in the future, addition to the strict interventions, more surveillance testing and effective testing strategies under conditions of limited test availability, such as contact tracing of the contacts and confirmed cases, will be helpful to curtail the pandemic in South Africa [12].

Although highly transmissible and poorly ascertained, the COVID-19 IFR is estimated as 0.06% taking account of unreported cases and deaths in South Africa, comparable to the estimates in other locations with similar low mortality rate based on serological data [19]. The low IFR may be due to the South African population being relatively young which lessens the fatal impact on general population to some extent [36]. Our estimates of the IFR of hospitalized cases are much higher than that for non-hospitalized cases, suggesting that the most severe cases may have been admitted to hospitals despite the relatively lack of the testing arrangements. The very low estimated IFR for the non-hospitalized cases also imply that the degree of under-reporting for death by the model is very low (0.24% by April 1, 2021), and likely to be affected by the same factors.

### Comparison of the models

The eSIR and the SAPHIRE model have been successfully applied to the data in India and Wuhan, China, separately [35, 13]. Although SAPHIRE model exhibits superior prediction performance on COVID-19 cases, the estimates of un-derlying paratemters and unascertained cases showed lack of robustness to the change of initial ascertainment rate *r*_0_. On the other hand, the eSIR model has the best estimation capability in terms of the ascertained cases but a relatively poor predictive capacity for capturing the change in the trend of the epidemic in time for neglecting some important clinical characteristics. The eSEIRD model also has a good fitting performance but a relatively poor prediction capacity. Table 2 also suggests that the estimates in eSEIRD model are robust estimated compared to the SAPHIRE model, probably an artifact of the Bayesian hierarchical model used.

### Strengths and Limitations

The key methodological innovation for the proposed method is revealed by a quick comparison between the schematic diagrams for eSEIRD model (Fig. 3) and SAPHIRE model (Fig. 2(b)). Broadly speaking, eSEIRD incorporates *π*(*t*), the transmission rate modifier as well as splits the ‘removed’ compartment into ‘recovered’ and ‘deaths’ while accounting for separate rates for ascertained, unascertained and hospitalized cases.

Despite the superior performance and robustness exhibited by the models examined here, there are some important limitations. First, the model assumptions were elicited from previous reports from other countries because of the lack of such information for South Africa, especially the fixed values for hyper-parameters. Though the estimation of parameters and prediction of infections seem to be robust to these assumptions to some extent, the inference and prediction would be much more convincing when based on accurate information specific to South Africa.

Second, the ascertained rate was assumed to follow the same distribution in a long time period in the eSEIRD model although in reality it might be time-varying depending on the accumulating knowledge and deployment of clinical resources for COVID-19, given the spatial variation within South Africa regarding the population density and movement, as well as regarding location of COVID-19 hotspots and hospital resources. Further, the population density is highly heterogeneous in different regions in South Africa with higher concentration near high-density economic hub cities, such as Cape Town and Durban. COVID-19 cases are also diversely spread. For instant, Gauteng Province is a very small, highly dense province with roughly 30% of total cases in the nation, and 49% of confirmed cases cluster in KwaZulu-Natal, Eastern Cape and Western Cape Province. In addition, the seroprevalence study also suggested that the prevalence may vary from city to city: 63% in the Eastern Cape, 52% in the KwaZulu Natal and 32% in the Northern Cape [38]. Without considering these heterogeneities and potential confounding factors in individual region, the conclusion on the national data might be biased. The burden of HIV and tuberculosis comorbidity, particularly among the less privileged socio-economic population, also adds to the complexity of analyzing the COVID-19 data from South Africa [5].

Third, in this paper we implicitly assumed that the recovered cases would not be infected again but it is still inconclusive based on extant research for COVID-19 [14]. It might lead to a resurgence if this assumption is not valid and the interventions are totally lifted. Thus, it might be necessary to conduct more national serological surveys on COVID-19 among the general population in South Africa to confirm the national, as well as provincial, seroprevalence. Such large-scale studies will also provide more powerful evidence to examine the evolving benefits of non-pharmaceutical interventions decisions and provide guidance to manage provincial level disparity.

Finally, from the early stage of this pandemic to now, there has been an explosive development in COVID-19 forecasting models but systematic comparison between the available models in terms of out-of-sample prediction and inference has been rare (see e.g. [11]) as are carefully done simulation studies where the ‘ground truth’ is known. Lack of simulation studies comparing the candidate methods is also a limiting feature of this paper, and we hope to pursue this in a future endeavor.

## Data Availability

The data of this study are openly available in the COVID-19 Data Repository by the Center for Systems Science and Engineering (CSSE) at Johns Hopkins University at https://github.com/CSSEGISandData/COVID-19.

## 5.1 Acknowledgements

This work was supported by grants from the National Science Foundation [grant numbers DMS-1712933 (to B.M.) and DMS-2015460 (to J.D.)] and from National Institute of Health – 1 R01 HG008773-01 (to B.M.). The first and second author (X.G. and B.M.) would also like to thank the Center for Precision Health Data Sciences at the University of Michigan School of Public Health, The University of Michigan Rogel Cancer Center and the Michigan Institute of Data Science for internal funding that supported this research.

