## Supplementary material for "COVID-19 PREDICTION IN SOUTH AFRICA: ESTIMATING THE UNASCERTAINED CASES- THE HIDDEN PART OF THE EPIDEMIOLOGICAL ICEBERG"

**S.1.** Method implementation.

**S.2.** Summary of different assumptions of the models.

**S.3.** Solution to $f_1(\boldsymbol{\theta_{t-1}}, \beta, \nu)$.

**S.4.** Solution to $f_2(\boldsymbol{X_{t-1}}, \beta, r, \kappa_1, \kappa_2)$.

**S.1.** Method implementation.

**Table S.1.1** Initial values and hyper-parameter choices.

| Parameters | Description | Settings |
|---|---|---|
| $\boldsymbol{\alpha}$ | Transmissibility ratio between unascertained and ascertained cases | Fixed as 0.55 |
| $\boldsymbol{\beta}$ | Transmission rate | Random |
| $\boldsymbol{D_e}$ | Latent period | Fixed as 2.9 days |
| $\boldsymbol{D_h}$ | Period from hospitalization to discharge or death | Fixed as 8.6 days |
| $\boldsymbol{D_i}$ | Symptomatic infectious period | Fixed as 2.9 days |
| $\boldsymbol{D_p}$ | Pre-symptomatic period | Fixed as 2.3 days |
| $\boldsymbol{D_q}$ | The period of ascertained cases from reporting to hospitalization | Fixed as 7 days |
| $\boldsymbol{\kappa_1}$ | Infection mortality rate for non-hospitalized cases | Random |
| $\boldsymbol{\kappa_2}$ | Infection mortality rate for hospitalized cases | Random |
| $\boldsymbol{N}$ | Population size in South Africa | Fixed as $N$=57,779,622 |
| $\boldsymbol{n}$ | The number of inbound or outbound individuals | Fixed as step function: $n$=0.0004$N$ on Mar 15-25; $n$=0 otherwise. |
| $\boldsymbol{\nu}$ | Removed rate | Random |
| $\boldsymbol{r}$ | Ascertained rate | Random |
| $\boldsymbol{R_e}$ | Effective reproduction number | Random |
| $\boldsymbol{R_0}$ | Basic reproduction number | Random |

**S.2.** Summary of different assumptions of the models.

| Model | eSIR | SAPHIRE | eSEIRD |
|---|---|---|---|
| **ODEs** | S,I,R | S,E,P,A,I,H,R | S,E,A,I,H,R,D |
| **Priors** | $\nu \sim LogN(-2.955, 0.910)$<br>$R_0 \sim LogN(1.14, 0.09)$<br>$\omega \sim Gamma(2, 0.0001)$<br>$\lambda^I \sim Gamma(2, 0.0001)$<br>$\lambda^R \sim Gamma(2, 0.0001)$ | $r_1 \sim Beta(10, 90)$<br>$\delta_1, \delta_2, \delta_3, \delta_4,$ and $\delta_5 \sim N(0,1)$<br>$\beta_1, \beta_2, \beta_3, \beta_4, \beta_5$ and $\beta_6 \sim Unif(0, 2)$ | $r_1, r_2, r_3, r_4, r_5$ *and* $r_6 \sim Beta(10, 90)$<br>$R_{01}, R_{02}, R_{03}, R_{04}, R_{05}$ and $R_{06} \sim LogN(1.14, 0.09)$<br>$\kappa_{11}, \kappa_{12}, \kappa_{13}, \kappa_{14}, \kappa_{15}$ and $\kappa_{16} \sim Beta(0.03, 2.93)$<br>$\kappa_{21}, \kappa_{22}, \kappa_{23}, \kappa_{24}, \kappa_{25}$ and $\kappa_{26} \sim Beta(0.44, 1.76)$ |
| **Solutions to the ODEs** | RK4 approximation | Pseudo likelihood | RK4 approximation |
| **Likelihood function components** | $Y_t^I/N \mid \boldsymbol{\theta_t}, \boldsymbol{\tau_1} \sim \mathrm{Beta}(\lambda_t^I \theta_t^I, \lambda^I(1-\theta_t^I))$<br>$Y_t^R/\mathrm{N} \mid \boldsymbol{\theta_t}, \boldsymbol{\tau_1} \sim \mathrm{Beta}(\lambda_t^R \theta_t^R, \lambda^R(1-\theta_t^R))$ | $Y_t^I \sim \mathrm{Poisson}(\lambda_t^I = \frac{rP_{t-1}}{D_p})$ | $Y_t^I \mid \boldsymbol{X_t}, \boldsymbol{\tau_3} \sim \mathrm{Poisson}(\lambda_t^I = \frac{rE_{t-1}}{D_e})$<br>$Y_t^R \mid \boldsymbol{X_t}, \boldsymbol{\tau_3} \sim \mathrm{Poisson}(\lambda_t^R = \frac{(1-\kappa_2)}{D_i} I_{t-1} + \frac{(1-\kappa_3)}{D_h} H_{t-1})$<br>$Y_t^D \mid \boldsymbol{X_t}, \boldsymbol{\tau_3} \sim \mathrm{Poisson}(\lambda_t^D = \frac{\kappa_2}{D_i} I_{t-1} + \frac{\kappa_3}{D_h} H_{t-1})$ |

**S.3.** Solution to $f_1(\boldsymbol{\theta_{t-1}}, \beta, \nu)$

$$k_t^{S1} = -\beta\pi(t)\theta_t^S\theta_t^I,$$

$$k_t^{S2} = -\beta\pi(t)(\theta_t^S + 0.5k_t^{S1})(\theta_t^I + 0.5k_t^{I1}),$$

$$k_t^{S3} = -\beta\pi(t)(\theta_t^S + 0.5k_t^{S2})(\theta_t^I + 0.5k_t^{I2}),$$

$$k_t^{S4} = -\beta\pi(t)(\theta_t^S + k_t^{S3})(\theta_t^I + k_t^{I3});$$

$$k_t^{I1} = \beta\pi(t)\theta_t^S\theta_t^I - \nu\theta_t^I,$$

$$k_t^{I2} = \beta\pi(t)(\theta_t^S + 0.5k_t^{S1})(\theta_t^I + 0.5k_t^{I1}) - \nu(\theta_t^I + 0.5k_t^{I1}),$$

$$k_t^{I3} = \beta\pi(t)(\theta_t^S + 0.5k_t^{S2})(\theta_t^I + 0.5k_t^{I2}) - \nu(\theta_t^I + 0.5k_t^{I2}),$$

$$k_t^{I4} = \beta\pi(t)(\theta_t^S + k_t^{S3})(\theta_t^I + k_t^{I3}) - \nu(\theta_t^I + k_t^{I3});$$

$$k_t^{R1} = \nu\theta_t^I,$$

$$k_t^{R2} = \nu(\theta_t^I + 0.5k_t^{I1}),$$

$$k_t^{R3} = \nu(\theta_t^I + 0.5k_t^{I2}),$$

$$k_t^{R4} = \nu(\theta_t^I + k_t^{I3});$$

**S.4.** Solution to $f_2(\boldsymbol{X_{t-1}}, \beta, r, \kappa_1, \kappa_2)$

$$k_t^{S1} = n - \frac{\beta\pi(t)S_t(I_t + \alpha A_t)}{N} - \frac{nS_t}{N},$$

$$k_t^{S2} = n - \frac{\beta\pi(t)(S_t + 0.5k_t^{S1})(I_t + 0.5k_t^{I1} + \alpha(A_t + 0.5k_t^{A1}))}{N} - \frac{n(S_t + 0.5k_t^{S1})}{N},$$

$$k_t^{S3} = n - \frac{\beta\pi(t)(S_t + 0.5k_t^{S2})(I_t + 0.5k_t^{I2} + \alpha(A_t + 0.5k_t^{A2}))}{N} - \frac{n(S_t + 0.5k_t^{S2})}{N},$$

$$k_t^{S4} = n - \frac{\beta\pi(t)(S_t + k_t^{S3})(I_t + k_t^{I3} + \alpha(A_t + k_t^{A3}))}{N} - \frac{n(S_t + k_t^{S3})}{N};$$

$$k_t^{E1} = \frac{\beta\pi(t)S_t(I_t + \alpha A_t)}{N} - \frac{E_t}{D_e + D_p} - \frac{nE_t}{N},$$

$$k_t^{E2} = \frac{\beta\pi(t)(S_t + 0.5k_t^{S1})(I_t + 0.5k_t^{I1} + \alpha(A_t + 0.5k_t^{A1}))}{N} - \frac{(E_t + 0.5k_t^{E1})}{D_e + D_p} - \frac{n(E_t + 0.5k_t^{E1})}{N},$$

$$k_t^{E3} = \frac{\beta\pi(t)(S_t + 0.5k_t^{S2})(I_t + 0.5k_t^{I2} + \alpha(A_t + 0.5k_t^{A2}))}{N} - \frac{(E_t + 0.5k_t^{E2})}{D_e + D_p} - \frac{n(E_t + 0.5k_t^{E2})}{N},$$

$$k_t^{E4} = \frac{\beta\pi(t)(S_t + k_t^{S3})(I_t + k_t^{I3} + \alpha(A_t + k_t^{A3}))}{N} - \frac{(E_t + k_t^{E3})}{D_e + D_p} - \frac{n(E_t + k_t^{E3})}{N};$$

$$k_t^{A1} = \frac{(1 - r)E_t}{D_e + D_p} - \frac{A_t}{D_i} - \frac{nA_t}{N},$$

$$k_t^{A2} = \frac{(1 - r)(E_t + 0.5k_t^{E1})}{D_e + D_p} - \frac{(A_t + 0.5k_t^{A1})}{D_i} - \frac{n(A_t + 0.5k_t^{A1})}{N},$$

$$k_t^{A3} = \frac{(1 - r)(E_t + 0.5k_t^{E2})}{D_e + D_p} - \frac{(A_t + 0.5k_t^{A2})}{D_i} - \frac{n(A_t + 0.5k_t^{A2})}{N},$$

$$k_t^{A4} = \frac{(1 - r)(E_t + k_t^{E3})}{D_e + D_p} - \frac{(A_t + k_t^{A3})}{D_i} - \frac{n(A_t + k_t^{A3})}{N};$$

$$k_t^{I1} = \frac{rE_t}{D_e + D_p} - \left(\frac{1}{D_i} + \frac{1}{D_q}\right)I_t,$$

$$k_t^{I2} = \frac{r(E_t + 0.5k_t^{E1})}{D_e + D_p} - (\frac{1}{D_i} + \frac{1}{D_q})(I_t + 0.5k_t^{I1}),$$

$$k_t^{I3} = \frac{r(E_t + 0.5k_t^{E2})}{D_e + D_p} - (\frac{1}{D_i} + \frac{1}{D_q})(I_t + 0.5k_t^{I2}),$$

$$k_t^{I4} = \frac{r(E_t + k_t^{E3})}{D_e + D_p} - (\frac{1}{D_i} + \frac{1}{D_q})(I_t + k_t^{I3});$$

$$k_t^{H1} = \frac{I_t}{D_q} - \frac{H_t}{D_h},$$

$$k_t^{H2} = \frac{(I_t + 0.5k_t^{I1})}{D_q} - \frac{(H_t + 0.5k_t^{H1})}{D_h},$$

$$k_t^{H3} = \frac{(I_t + 0.5k_t^{I2})}{D_q} - \frac{(H_t + 0.5k_t^{H2})}{D_h},$$

$$k_t^{H4} = \frac{(I_t + k_t^{I3})}{D_q} - \frac{(H_t + k_t^{H3})}{D_h};$$

$$k_t^{R1} = (\frac{(1-\kappa_1)I_t}{D_i} + \frac{(1-\kappa_1)A_t}{D_i}) + \frac{(1-\kappa_2)H_t}{D_h} - \frac{nR_t}{N},$$

$$k_t^{R2} = (\frac{(1-\kappa_1)(I_t + 0.5k_t^{I1})}{D_i} + \frac{(1-\kappa_1)(A_t + 0.5k_t^{A1})}{D_i}) + \frac{(1-\kappa_2)(H_t + 0.5k_t^{H1})}{D_h} - \frac{n(R_t + 0.5k_t^{R1})}{N},$$

$$k_t^{R3} = (\frac{(1-\kappa_1)(I_t + 0.5k_t^{I2})}{D_i} + \frac{(1-\kappa_1)(A_t + 0.5k_t^{A2})}{D_i}) + \frac{(1-\kappa_2)(H_t + 0.5k_t^{H2})}{D_h} - \frac{n(R_t + 0.5k_t^{R2})}{N},$$

$$k_t^{R4} = (\frac{(1-\kappa_1)(I_t + k_t^{I3})}{D_i} + \frac{(1-\kappa_1)(A_t + k_t^{A3})}{D_i}) + \frac{(1-\kappa_2)(H_t + k_t^{H3})}{D_h} - \frac{n(R_t + k_t^{R3})}{N};$$

$$k_t^{D1} = (\frac{\kappa_1 I_t}{D_i} + \frac{\kappa_1 A_t}{D_i}) + \frac{\kappa_2 H_t}{D_h},$$

$$k_t^{D2} = (\frac{\kappa_1(I_t + 0.5k_t^{I1})}{D_i} + \frac{\kappa_1(A_t + 0.5k_t^{A1})}{D_i}) + \frac{\kappa_2(H_t + 0.5k_t^{H1})}{D_h},$$

$$k_t^{D3} = (\frac{\kappa_1(I_t + 0.5k_t^{I2})}{D_i} + \frac{\kappa_1(A_t + 0.5k_t^{A2})}{D_i}) + \frac{\kappa_2(H_t + 0.5k_t^{H2})}{D_h},$$

$$k_t^{D4} = \left(\frac{\kappa_1(I_t + k_t^{I3})}{D_i} + \frac{\kappa_1(A_t + k_t^{A3})}{D_i}\right) + \frac{\kappa_2(H_t + k_t^{H3})}{D_h}.$$

9. Song PX, Wang L, Zhou Y, et al. An epidemiological forecast model and software assessing

interventions on COVID-19 epidemic in China. *MedRxiv*. Published online 2020.

10. Hao X, Cheng S, Wu D, Wu T, Lin X, Wang C. Reconstruction of the full transmission dynamics of COVID-19 in Wuhan. *Nature*. Published online July 16, 2020:1-5. doi:10.1038/s41586-020-2554-8

11. Lau H, Khosrawipour T, Kocbach P, Ichii H, Bania J, Khosrawipour V. Evaluating the massive underreporting and undertesting of COVID-19 cases in multiple global epicenters. *Pulmonology*. Published online 2020.

12. Bhattacharyya R, Bhaduri R, Kundu R, Salvatore M, Mukherjee B. Reconciling epidemiological models with misclassified case-counts for SARS-CoV-2 with seroprevalence surveys: A case study in Delhi, India. *medRxiv*. Published online 2020.

13. Butcher JC. *Numerical Methods for Ordinary Differential Equations*. 2nd ed. Wiley; 2008.

14. Ray D, Salvatore M, Bhattacharyya R, et al. Predictions, Role of Interventions and Effects of a Historic National Lockdown in India's Response to the the COVID-19 Pandemic: Data Science Call to Arms. *Harvard Data Science Review*. Published online June 9, 2020. doi:10.1162/99608f92.60e08ed5

15. Rahmandad H, Lim TY, Sterman J. Estimating COVID-19 under-reporting across 86 nations: implications for projections and control. *medRxiv*. Published online August 3, 2020:2020.06.24.20139451. doi:10.1101/2020.06.24.20139451

16. University of Oxford. CORONAVIRUS GOVERNMENT RESPONSE TRACKER. https://www.bsg.ox.ac.uk/research/research-projects/coronavirus-government-response-tracker
